# Expansion of clinical genetic testing since the completion of the human genome project

**DOI:** 10.1101/2024.10.17.24315685

**Authors:** Lisa Bastarache, Rory J. Tinker, Bryce A. Schuler, Lucas Richter, John A. Phillips, William W. Stead, Gillian Hooker, Josh F. Peterson, Douglas M. Ruderfer

## Abstract

The sequencing of the first human genome led to expectations of the widespread use of genetics in medicine. However, assessing the true impact of genetic testing on clinical practice is challenging due to the lack of integration in the electronic health record (EHR). We extracted clinical genetic tests from the EHRs of over 1.8 million patients seen at Vanderbilt University Medical Center from 2002 to 2022, using both automated and manual methods. Using these data, we quantified the extent of clinical genetic testing in healthcare and described how testing patterns have changed over time, including utilization rate, test comprehensiveness, diagnoses made, and the number of variants of uncertain significance (VUS) returned.

We also assessed genetic testing rates across medical specialties and introduce a measure – the genetic attributed fraction (GAF) – to compute the proportion of observed phenotypes attributable to a genetic diagnosis. We identified 104,392 tests, 32% of which were only reported in unstructured text, and 19,032 molecularly confirmed diagnoses or risk factors. The proportion of patients genetic testing recorded in their EHRs from 1.0% in 2002 to 6.1% in 2022, and testing became more comprehensive with the growing use of multigene panels. This corresponded with a substantial increase in the variety of diseases diagnosed with genetic testing, from 51 unique diseases in 2002 to 509 in 2022, alongside a growing number of VUS.

The phenome-wide GAF for 6,505,620 diagnoses made in 2022 was 0.46%, with 74 phenotypes having a GAF greater than 5%, including pancreatic insufficiency (67%), chorea (64%), atrial septal defect (24%), Microcephaly (17%), paraganglioma (17%), and ovarian cancer (6.8%). Our study provides a comprehensive quantification of the increasing role of genetic testing at a major academic medical institution. These results demonstrate the now pervasive use of genetic testing across diverse medical contexts and its growing utility in explaining observed medical phenome.

## Introduction

Over the last 20 years, our ability to interrogate the human genome has become more robust, efficient, and cost-effective, resulting in more opportunities to use genetic testing for clinical purposes.^1–3^ However, informatics solutions to integrate these tests into electronic health records (EHRs) have lagged behind.^4^ Researchers have long envisioned a system where genetic test results are stored in a searchable format and easily flow between systems, similar to other laboratory test results.^5^ In reality, genetic test results are often stored in the EHR as scanned documents that are not machine-readable and hidden from easy accounting.^6–8^

The lack of EHR integration makes it challenging to measure the impact of genetic testing in healthcare. Most studies of genetic testing utilization have focused on specific clinical contexts or testing indications^11,12^ and often rely on imperfect datasets, such as coded billing data, which lack specificity and are inconsistently applied, or data from testing laboratories which lack medical context and longitudinality.^9,10^ Despite these limitations, prior studies have highlighted key trends, including the increased use and enhanced diagnostic capability of genetic testing within certain clinical populations,^11,13–15^ as well as current challenges related to inclusive testing results and inefficiencies of the testing process.^16–19^ However, a full accounting of how genetic testing has been adopted across subspecialities and indications has not yet been performed.

To address this gap, we generated a clinical genetics database (CGdb) by extracting genetic testing results from the EHRs of 1.86 million patients at the Vanderbilt University Medical Center from 2002 to 2022. Using both automated parsing and manual chart review, we uncovered a substantial amount of genetic testing hidden in unstructured text of the EHR. Our study tracks the cumulative growth of clinical genetic testing and illustrates the complexity of extracting such information given the current lack of structure. We also analyze the evolution of genetic testing over the past two decades, noting a shift towards more comprehensive testing. Additionally, we catalog diagnoses made through genetic testing, including the many rare conditions that are enriched in a tertiary healthcare population, and quantify the accumulation of uncertain test results. Finally, we assess patient exposure to genetic testing across specialties and measure the morbidity explained by genetic diagnoses across phenotypes. Our results demonstrate that genetic testing is now pervasive in medicine, and we provide examples of the value of structured clinical genetic testing data for both operational and research purposes.

## Methods

### Study population

Our study focused on patients seen at Vanderbilt University Medical Center (VUMC) from January 1, 2002, to December 31, 2022. All data were derived from the research derivative (RD), an image of the structured and textual data elements of the EHR, including demographics, claims data (e.g., International Classification of Disease [ICD] codes), clinical notes, problem lists, pathology reports, visit information, vital signs, and laboratory measures. The cohort included individuals with at least three encounters spaced a minimum of 90 days apart or those who were born at VUMC. To avoid an influx of thinly phenotyped patients during the COVID pandemic, we excluded ICDs for vaccination, testing, or prophylactic treatment of infectious disease. Outpatient visit dates and clinic type/subspecialty information were obtained using the care site identifier linked to each patient’s visit. Outpatient visits included office visits, outpatient evaluations, and telemedicine visits. Race and gender were extracted from the demographics table in the EHR. State of residence and insurance type were recorded based on the most recent entry for each patient. All ICD codes were mapped to phecodeX.^20^ See Supplementary Table 1 for data element definitions.

**Table 1.** The prevalence of genetic disorders as estimated by OphraNet and measured in the CGdb. The table includes ten most common genetic diagnoses in CGdb with available prevalence data available from OrphaNet.

| OrphaCode | Disease | # affected in CGdb | Prevalence estimates (per 100,000) |  | Estimated population biobank size |
| --- | --- | --- | --- | --- | --- |
|  |  |  | OrphaNet | CGdb |  |
| 870 | Down syndrome | 1046 | 57 | 56.95 | 1,835,088 |
| 586 | Cystic fibrosis | 962 | 11.13 | 52.38 | 8,641,831 |
| 399 | Huntington disease | 335 | 2.7 | 18.24 | 12,407,407 |
| 881 | Turner syndrome | 325 | 5.5 | 17.69 | 5,909,091 |
| 567 | DiGeorge syndrome | 286 | 37.5 | 15.57 | 762,667 |
| 363700 | Neurofibromatosis, type 1 | 234 | 21.3 | 12.74 | 1,098,592 |
| 98896 | Duchenne muscular dystrophy | 195 | 2.8 | 10.62 | 6,964,286 |
| 558 | Marfan syndrome | 167 | 15 | 9.09 | 1,113,333 |
| 98878 | Hemophilia A | 103 | 4.85 | 5.61 | 2,123,711 |
| 273 | Myotonic dystrophy 1 | 97 | 12.5 | 5.28 | 776,000 |

### dentifying and processing genetic test results

Our goal was to comprehensively extract and curate germline genetic tests from the EHR, including single-variant or single-gene tests, multi-gene panels (tests that analyze two or more genes), exome and genome sequencing (ES/GS), nucleotide repeat expansions, chromosomal microarrays (CMA), and karyotypes. We also included methylation tests for syndromic disorders (e.g., Beckwith-Wiedemann syndrome, Angelman syndrome). We did not attempt to extract pharmacogenetic testing, somatic tumor testing, or circulating fetal DNA tests.

Genetic tests were recorded in the EHR in both structured and unstructured formats. Tests run by the local pathology laboratory were recorded in the EHR as templated pathology reports, including the majority of CMAs, karyotypes, and single gene/variant tests for *HFE, F2, MTHFR*, etc. Tests from external laboratories were typically linked to the EHR as non-searchable PDF reports; however clinicians also recorded these results as templates embedded in clinical notes. These templates were most common in clinics with a high volume of testing, including medical genetics and the hereditary cancer clinic. Tests recorded as pathology reports or templates were identified and processed using regular expressions and custom parsers. Tests not represented in any structured format were more difficult to process. To locate these tests, we indexed the EHRs in our cohort for 4,207 gene names from the Online Mendelian Inheritance in Man (OMIM) (Supplementary File 1) and names of genetic testing laboratories. We extracted a 200-character window around each gene name and filtered these mentions for false positives using keywords. Notes with gene names underwent human review, and positive mentions (i.e., those referring to germline testing for the patient) were curated using a terminal-based interface developed for this project. Data elements curated included the test type (e.g., single gene, multigene panel), indication (e.g., diagnostic, cascade testing, carrier screening), and variants returned from testing. PDFs were reviewed to cross-check information recorded in the notes and retrieve information missing from clinical notes. Each test instance was annotated by the test type, name, date, and any variants returned. Variants were annotated based on the interpretation provided by the clinical testing laboratory.

Test results were annotated as diagnostic if they detected pathogenic variant(s) that were consistent with a diagnosis documented in the EHR. Diagnostic test results were linked to unique disease identifiers from OMIM or Orphanet if OMIM lacked a specific disease identifier.^21,22^ Twenty-seven diagnoses were defined as risk factors as they contributed to a multifactorial disorder with both genetic and environmental components (e.g., Factor V Leiden thrombophilia, MIM:188055). (Supplementary file 2) Tests were labeled as “carrier” if they returned one or more pathogenic or likely pathogenic variants linked to an autosomal recessive gene, “inconclusive” if they returned one or more variants of uncertain significance (VUS), and “negative” if they returned no variants. A clinical geneticist grouped genetic panel tests into broad disease categories (e.g., cancer, endocrinology, cardiology).

### Linking diseases and genes to phenotypes

We linked genes and genetic diagnoses to phecodes, an ICD-based method than capture of diagnoses, signs and symptoms across the medical phenome.^23^ To do so, we used the human phenotype ontology (HPO) annotations for rare disease (version 2024-04-24) that link genetic disease and genes to phenotypes encoded as HPO terms.^24^ We developed an HPO to phecodeX map such the phenotypic manifestations of each disease and gene could be described as a set of phecodes (per methods described in prior publications).^25,26^ To link diagnoses and test results to medical subspecialties, we mapped 12 of HPO high-level phenotype categories to corresponding subspecialties (e.g. Abnormality of the nervous system->Ophthalmology; Neoplasm->Oncology). (Supplementary files 3-5)

### Descriptive statistics for tests and diagnoses

To assess changes in genetic testing utilization over time, we summarized the cumulative and yearly counts for the number of tests, diagnoses, and variants returned, stratified by test type and/or indication. We also summarized the test types and panel disease categories by diagnostic yield (i.e. the percentage of tests that returned a diagnostic result), as well as the fraction of tests that returned carrier status (i.e. a single pathogenic variant for an autosomal recessive disease), uncertain variants, or no results (negative).

### Assessing testing rates across subspecialty clinics

We assessed the percentage of patients exposed to genetic testing over time, with a denominator as the number of patients who were seen at VUMC for each target year and the numerator as the number of patients who had genetic testing in their EHR. We stratified the percent tested measure across all outpatient clinics that saw at least 200 patients in our cohort in 2022. For 12 subspecialties, we further assessed the relevance of the genetic testing present in patients records by linking their diagnoses and results to subspecialties.

### Computing the genetic attributed fraction (GAF)

Genetic diagnoses can explain a wide range of signs and symptoms. For instance, a diagnosis of cystic fibrosis clarifies why a patient may experience bronchiectasis and pancreatitis, while hereditary TTR amyloidosis accounts for a patient’s heart failure and neuropathy. We developed a metric called the genetic attributed fraction (GAF) to measure the proportion of a phenotype that can be linked to a genetic diagnosis. The GAF is calculated as follows: the denominator is the total number of patients with a specific phenotype (cases), while the numerator is the number of these cases that have a genetic diagnosis associated with that to that phenotype (diagnosed cases). We defined cases as individuals with a specific phenotype code (phecodes) recorded on at least two distinct dates within the target year. We linked phecodes from patient EHRs to genetic diagnoses using the HPO to phecode map described earlier. We computed phenome-wide GAF for all phenotypes observed in 2022 (excluding phecodes in the genetic diagnosis chapter) as well as for individual phenotypes. Additionally, we assessed the change in GAF from 2002 to 2022 for 25 phecodes with at least 100 cases, a GAF>5%, and linkage to more than one genetic disease. Only one phecode per parent code was included. We similarly linked non-diagnostic test results to the target phenotype using the HPO to gene mapping.

## Results

### Cohort characteristics

Our cohort included 1,836,752 patients with a median age of 36 at their last encounter at VUMC (interquartile range [IQR] 16–60) with a median of 12 encounters (IQR 6–28) and a median of 5.0 years (IQR 1.7-10.9) between the first and last encounter. 5.1% of patients were born at VUMC; and 33.9% had at least one encounter as a pediatric patient (age <18). The cohort was linked to 314,313,905 clinical notes from the EHR. A full description of cohort demographics can be found in Table S2.

### The EHR Contains a Significant and Expanding Repository of Clinical Genetic Tests and Diagnoses

Collectively, information relating to genetic tests, results, and diagnoses were compiled into a resource we call the clinical genetic database (CGdb), which includes a cumulative total of 104,392 genetic tests for 77,033 patients (Figure 1). Overall, multi-gene panels were the most common test type (26.0%), followed by single genes (23.1%), single variant (17.3%) and CMA (14.3%), karyotype (9.7%), repeat expansion (7.1%), ES/GS (1.3%), and methylation tests (1.1%). By indication, most tests were for diagnostic purposes (85.8%); carrier screening and familial variant testing accounted for 9.6% and 4.4%, respectively. In terms of results, the majority of tests were negative (63.1%); 15.7% were diagnostic, 2.7% returned a risk factor, 5.9% returned carrier status, and 10.1% returned one or more VUS and no diagnosis. (Table S3).

**Figure 1.**
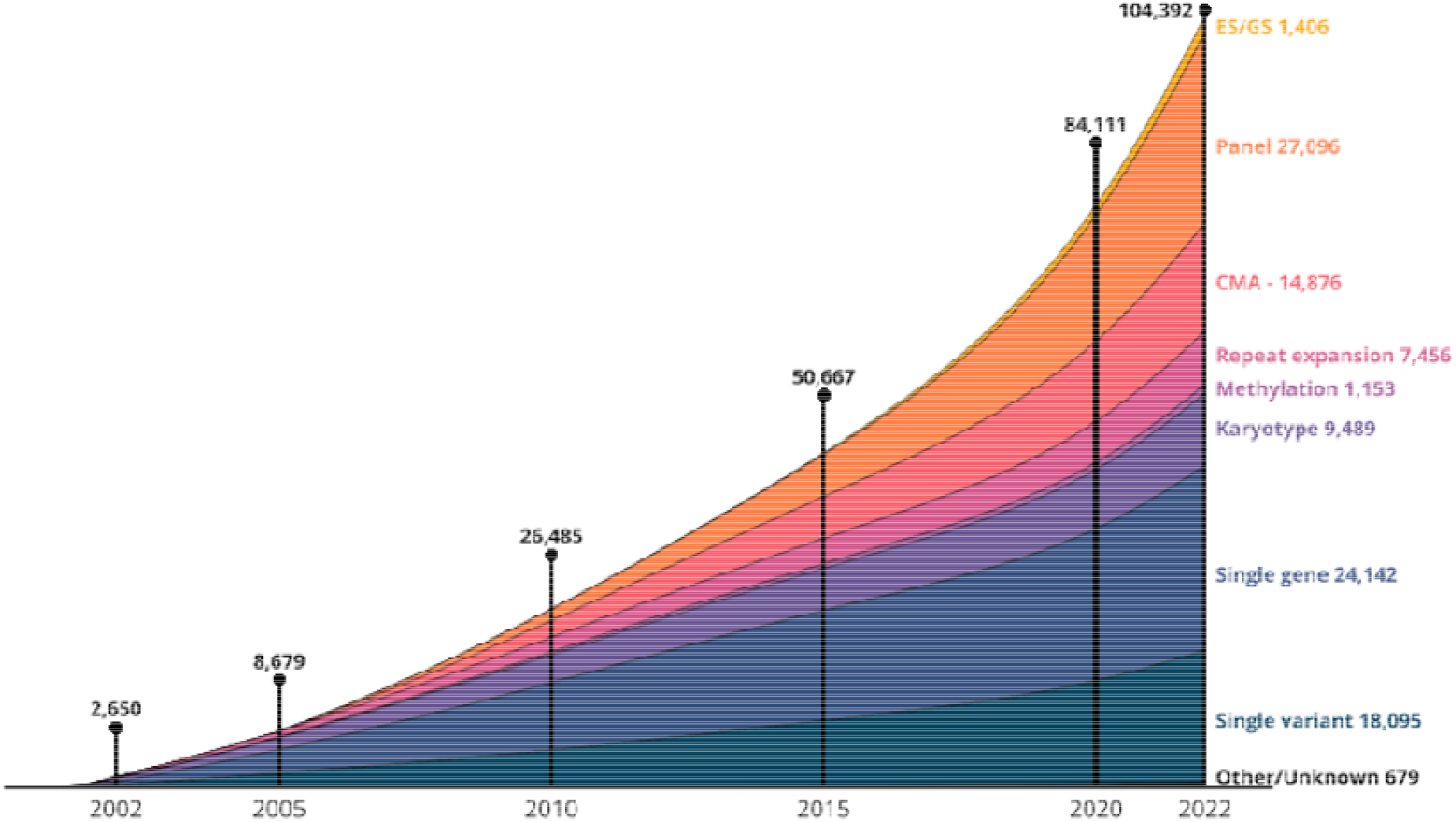
The cumulative growth of genetic testing recorded in the EHR. The cumulative number of tests by year, stratified by test type. Cumulative counts of tests are shown at years 2002, 2005, 2010, 2015, and 2022, and cumulative totals for each test type are labeled on the right side of the figure.

Overall, the CGdb comprises 19,032 diagnoses and risk factors for 18,476 patients; 533 patients (2.8%) had more than one diagnosis, similar to rates reported in other studies.^27,28^ The most common genetic diagnoses were Trisomy 21 (n=1,046; MIM:190685), Factor V Leiden thrombophilia (n=993; MIM:188055), cystic fibrosis (n=962; MIM:219700), and *BRCA2*-related familial breast-ovarian cancer (n=626; MIM:612555). Most diseases were rare: 85% (n=1,330) were diagnosed in fewer than 10 patients, and 43% (n=673) were diagnosed in a single patient. Collectively, the CGdb includes patients with diagnoses 1,564 different diseases, including 1,502 diseases annotated with OMIM identifiers, which constitutes 20% of the 7,528 disorders currently cataloged in OMIM (Tables S4 and S5). Based on Orphanet’s 2023 estimates of disease prevalence, ^29,30^ a population-based biobank would need over 8.6 million individuals to have the equivalent number cystic fibrosis patients as found in the CGdb, and over 12 million for Huntington disease. (Table 1).

### No consistent convention for documenting genetic testing or diagnosis in the EHR

Genetic testing and diagnostic information were scattered throughout the EHR in multiple formats and building the CGdb necessitated both custom automated extraction methods and substantial manual review. We identified 65,836 structured reports generated by the pathology lab at VUMC, 80.8% of which were for CMAs, karyotypes, and single gene or variant tests. The remaining 38,556 tests were embedded in clinical notes, including 5,881 tests recorded in templated language pasted within the body of a note and 32,745 tests that lacked any consistent formatting. To locate genetic testing mentioned in free text only, we indexed all notes for gene mentions, yielding 3,526,486 mentions for 432,581 patients and 3,994 unique genes. Only 10% of gene mentions indicated results of a genetic test. False positive mentions included synonymy for short gene names (e.g., ESPN, OTC), mentions of genetic variants found in family members, discussions of possible genetic testing, and somatic variants. Overall, 36.9% of tests were recorded in the clinic notes with no structure, including most multi-gene panels and ES/GS (81.6% and 64.2%, respectively). The fraction of tests reported in free text only increased over time, from 14.8% in 2002 to 48.9% in 2022 (Figure S1).

Genetic tests originated from 112 different laboratories, accounting for 32,218 of tests in the CGdb. Eight external labs contributed more than 1,000 tests each, accounting for 25.1% of tests overall. 103 additional external laboratories contributed an additional 6,029 tests.

### Growth of genetic testing over 21 years

An increasing percentage of the cohort received genetic testing over the course of the study period. Six-fold more patients had genetic testing recorded in their EHR in 2022 (6.1%) compared to in 2002 (1.0%) as illustrated in Figure S2. The number of new tests increased over time except for 2020 where there was a notable decrease in genetic testing, likely due to healthcare disruptions during the COVID-19 pandemic (Figure S3). In 2002, the percentage of patients with a molecularly confirmed diagnosis or genetic risk factor was 0.2%, increasing to 1.4% in 2022.

### A shift towards more comprehensive testing increased the variety of diagnoses made with genetic testing

In 2002, the most common test types were single gene (33.0%) and single variant tests (28.0%). Multigene panels grew in popularity over the study period. In 2015, multigene panels overtook single gene tests as the most popular test type and accounted for 46.3% of testing in 2022 (Figure 2A). The first ES/GS was reported in 2011 with 349 new ES/GS reported in 2022 surpassing the number of karyotypes that year (n=314).

**Figure 2.**
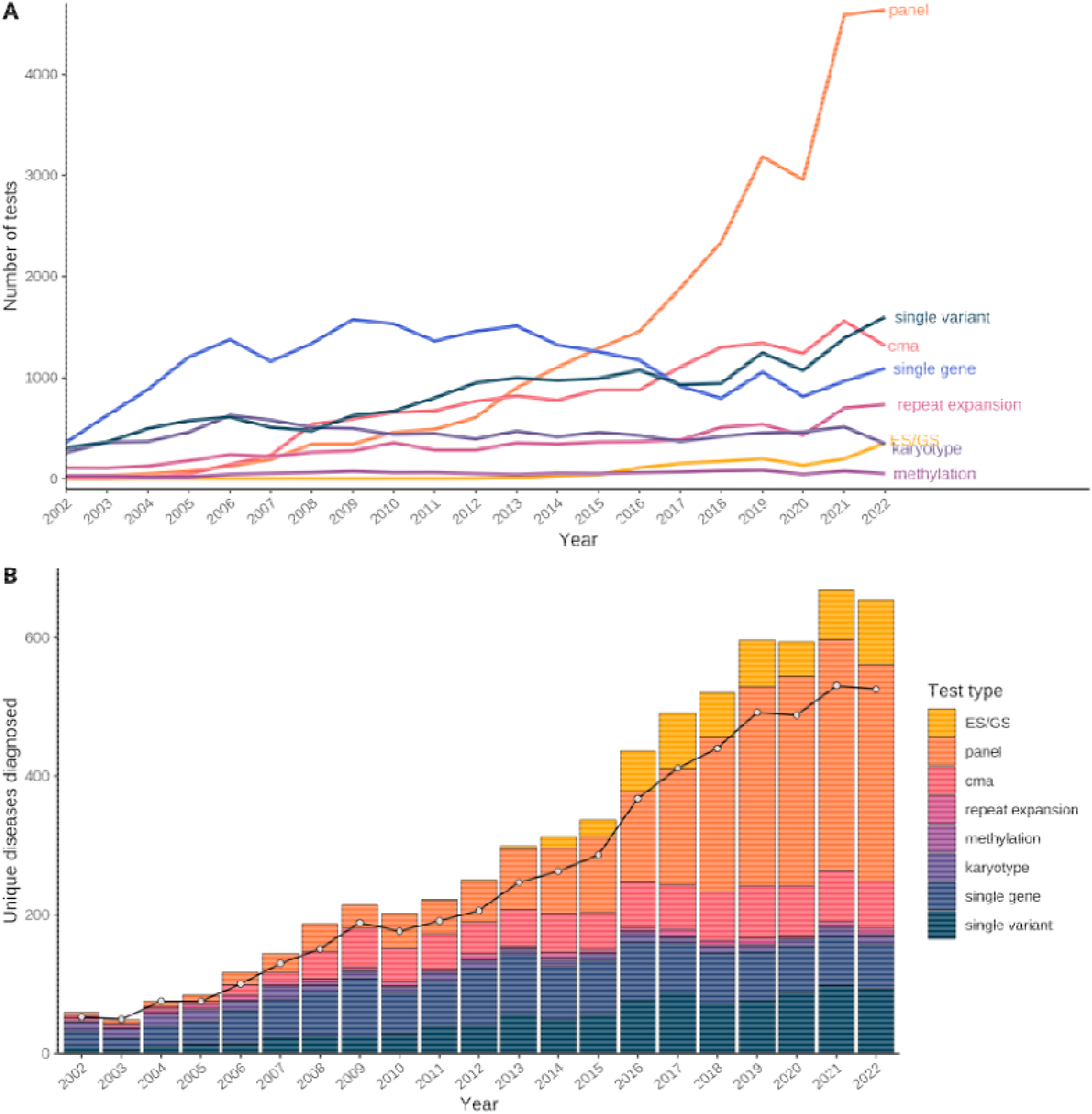
Trend towards more comprehensive tests and unique diagnoses. **(A)** The number of new genetic tests per year, stratified by test type. **(B)** The unique number of diagnoses per year made with different test types. Some diseases were diagnosed with different test types in the same year; the black line represents the overall number of unique diagnoses made in that year, regardless of test type.

As genetic testing became more comprehensive, the number of different diseases diagnosed with genetic testing increased, from 51 in 2002 to 509 in 2022 (Figure 2B). Multi-gene panels diagnosed 984 unique genetic diseases, or 63% of all diseases observed. Despite making up only 1.5% of tests overall, ES/GS yielded diagnostic results for 410 different diseases, 168 of which were not diagnosed with any other test type.

### More comprehensive testing associated with increased diagnostic yield as well as uncertainty

Overall, 28,646 pathogenic variants and 25,623 VUS were returned from genetic testing. While the cumulative number of pathogenic variants was greater than VUS, by 2018 VUS began accumulating at a faster rate than pathogenic variants, with 1.7 VUS per pathogenic variant by 2022 (Figure S4). The increase in the rate of VUS corresponded with the increase in multigene panels and ES/GS testing, which returned an average of 2.6 and 1.9 VUS per pathogenic variant, respectively. While ES/GS was the most likely test to return a diagnosis (3 .6%), they were also the most likely to return inconclusive results (29.8%, Figure S5). We also observed differences in the diagnostic yield across panel test categories, with tests for vision disorders having the highest diagnostic yield (49.5%) and obesity the lowest (1.5%, Figure S6).

### Genetic testing is pervasive across medical specialties

To determine the pervasiveness of genetic testing, we assessed the genetic testing rates among patients who visited various outpatient clinics. 6.5% of patients with an outpatient visit in 2022 had one or more genetic test in their EHR. The percentage of patients tested varied by clinic type, ranging from 1.1% for student health to 80.5% for medical genetics (Figure S7; Table S6). Because not all genetic tests are relevant to every subspecialist, we further analyzed the relevance of genetic testing across 12 medical specialties. For patients with outpatient visits with these subspecialties, an average of 11.9% (SD 4.4%) had testing in their EHR; 2.6% (SD 1.7%) had a relevant diagnosis, 2.8% (SD 1.0%) had a relevant inconclusive test, and 0.7% (SD 1.5%) had a relevant negative test (Figure 3).

**Figure 3.**
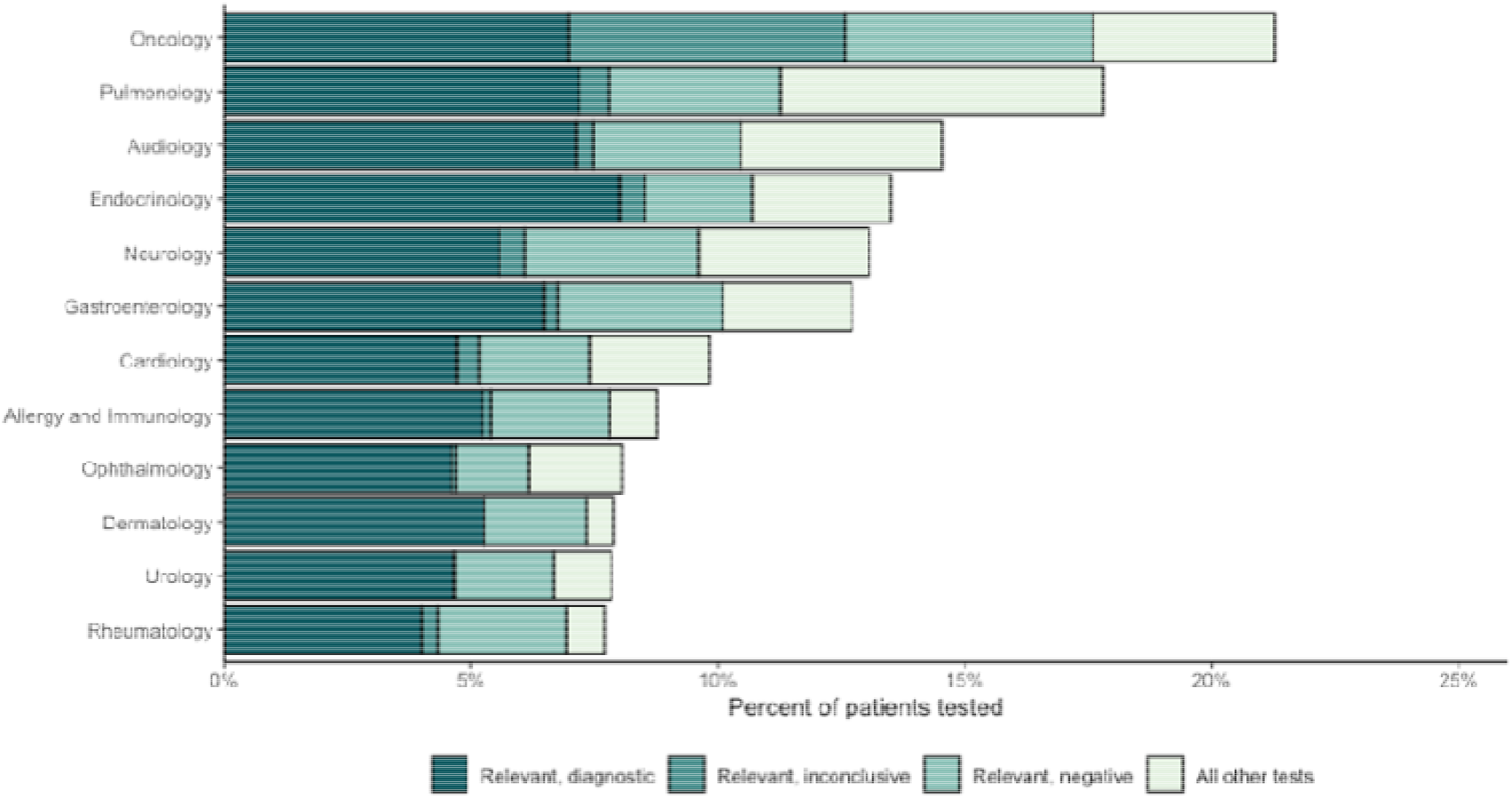
Percentage of patients with genetic testing in their EHR, by clinic specialty. The bars show the fraction of patients with genetic testing in the EHR, stratified by clinical specialty. Only outpatient or telemedicine visits in 2022 were included. Genetic tests were deemed relevant to the subspecialty if they were linked with a phenotype related to the subspecialty (e.g. Breast cancer is relevant to oncology; hypothyroidism is relevant to endocrinology). The definition of the test categories are as follows: Relevant, diagnostic includes tests diagnostic for a condition linked to the subspecialty; Relevant, inconclusive includes non-diagnostic tests that returned a VUS or carrier status linked to the subspecialty; Relevant negative includes all other unlinked tests.

### A substantial and increasing proportion of the observed medical phenome can be attributed to genetic diagnoses

Finally, we assessed the proportion of diagnoses in our population attributable to a diagnosed genetic disease using the genetic attributable fraction (GAF, see Methods). In 2022 patients received 6,462,640 diagnoses as encoded by 3,201 unique phecodes of which 29,545 were linked to a genetic diagnosis, resulting in a phenome-wide GAF of 0.46%. The GAF increased over time across a variety of unrelated phenotypes (Figure 4). Of the 2,049 phecodes with at least 100 cases, 788 had a GAF>0%. The phecodes with the highest GAF were Exocrine pancreatic insufficiency (67.1%), Chorea (63.5%), and Long QT syndrome (56.4%). Developmental delay had a GAF of 6.2% (659 of 10,704 patients) and was linked to 301 different genetic diagnoses, the largest number of any phenotype. Nine neoplasm phecodes had a GAF>4%, including paragangliomas (16.6%), ovarian cancer (8.6%), and breast cancer (4.8%, Figure S8)

**Figure 4.**
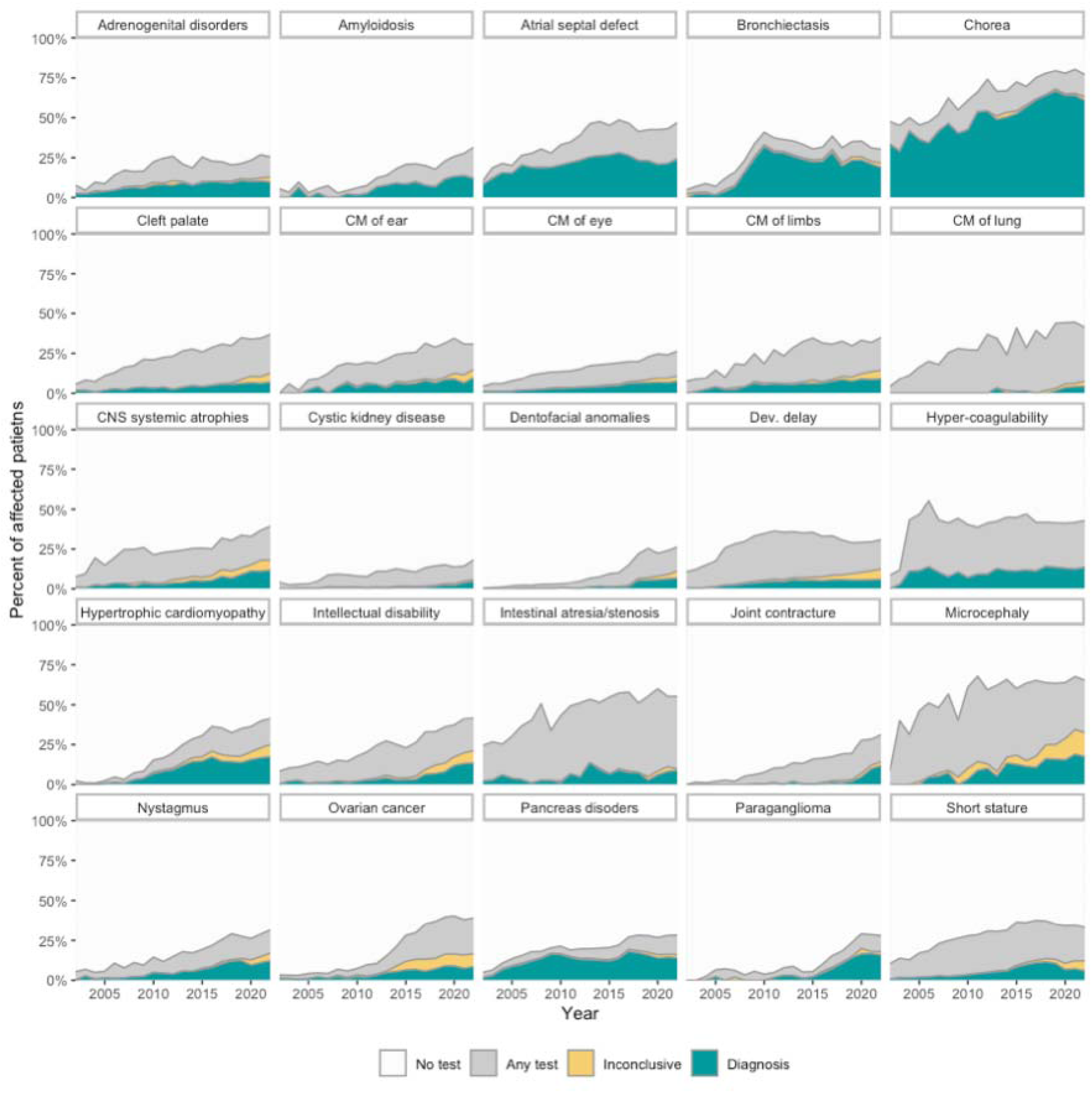
The percentage of phenotypes attributable to genetic diagnoses. Each square represents data for a single phenotype with the GAF (green), the fraction of patients with an inconclusive test (yellow), and the fraction of patients with genetic testing not linked to the phenotype (grey). Phecode labels were abbreviated for space. Abbreviations CM: Congenital malformation CNS: Central nervous system

## Discussion

By comprehensively curating genetic testing from the EHR, we were able to characterize its use and value across medical specialties and over time. Our findings were broadly consistent with previous studies, showing an increase in clinical genetic testing over the last 20 years, a trend towards more comprehensive testing, a corresponding increase in the total number and diversity of diagnoses, and an increased number of inconclusive test results. Unlike in prior studies, here we show how these trends manifest across test types and indications for an entire medial population.

Our study demonstrates the increasing role of genetic testing in clinical medicine. The rate clinical genetic testing increased year over year from 2002 to 2022, consistent with prior studies that looked at genetic testing for particular populations including hereditary cancer screening and preimplantation screening.^11,13^ In contrast to prior studies, we provide information on testing across the entire medical enterprise that allows for a more comprehensive understanding of how wide-spread clinical genetic testing has become since the completion of the human genome project. By 2022, nearly 1 in 16 patients had received some form of genetic testing during their clinical care. Importantly, this number continues to grow and while growth has occurred in nearly every area of medicine. This includes over 20% of patients that have been seen in oncology, similar to prior estimates in patients with breast or ovarian cancer,^31^ but less than 7% for patients seeing urology or rheumatology. A growing number of clinicians are faced with the challenge of managing patients with genetic testing while, at the same time, these tests are becoming more complex to interpret and utilize.

Our results also show the real-world impact of the increasing diagnostic power of genetic testing. Over the course of the study, the number of unique diseases diagnosed with genetic testing each year increased 10-fold – from 51 in 2002 to 509 in 2022 – representing great progress in the ability to detect a wide variety of diseases with genetic testing. These genetic diagnoses explained a significant and growing fraction of the observed phenome. Among the nearly 6.5 million phenotypes observed in 2022, 0.46% could be linked to a genetic diagnosis. This measure is one example of how EHR-linked genetic test data can be used to track the utility of genetic testing at the population level and measure the effect of interventions.

Our findings also reflect the known tension between the improved diagnostic resolution of comprehensive testing and the rise in VUS.^12,32,33^ We found that the highest rate of uncertain tests came from ES/GS followed by panel testing. Here our results contrast with a recent study of over 1.5 million genetic test results from 19 clinical laboratories, which found that the rate on inconclusive tests was higher for multi-gene panels compared with ES/GS.^34^ These differences likely reflect differences in variant reporting guidelines and local testing practices, highlighting the importance of assessing the utility of genetic testing in a variety of real-world settings. The shift toward more comprehensive testing is driving a growing demand for improved tools to support variant annotation and interpretation^35,36^ and calls to reexamine reporting practices.^37^

In addition to these findings, our work broadly demonstrates the value of aggregating genetic data across a medical system for two distinct purposes. First, if collected systematically, clinical genetic testing data may be used to identify emerging trends in the fast-evolving field of genetic testing and reveal inefficiencies in current practice. For example, curating where genetic testing is being performed across the hospital could provide opportunities to assess where testing might be expanded to increase diagnostic yield and reduce diagnostic delay.^38^ These data can be used to guide more effective use of genetic testing, enabling studies of medical practice and promoting a virtuous cycle of health improvement as envisioned by proponents of the learning healthcare system.^39^ Second, our study reveals that the EHR of a single academic medical center contain a wealth of genetic testing and diagnoses that could be used to improve knowledge and treatment of genetic disease. Prior studies show the value of EHR-based cohorts to increase our understanding of the phenotypic manifestations and natural history of rare disorders,^40^ improve our understanding of the epidemiology of these conditions,^41^ identify patients for clinical trials,^42^ and train machine learning algorithms to facilitate early detection.^43^ An EHR-linked CGdb constitutes a kind of biobank, one that is highly enriched for rare genetic disease and which continues to grow over time. With nearly 20,000 patients with a genetic diagnosis across over 1,500 diseases at VUMC alone a resource across multiple medical centers, a diagnosed disease network, could revolutionize the study of rare diseases.

A major challenge to curating clinical genetic testing is the poor organization in the EHR, necessitating substantial efforts to extract these data. More than one-third of tests were not linked to structured reports but were instead embedded within narrative text. The proportion of these unstructured tests increased over the study period, driven by the growing use of send-out tests which originated from more that 100 different external laboratories. The lack of searchable, machine-readable test results is a well-known and widespread problem.^44^ Standards have been developed to facilitate EHR integration,^45^ but progress has been slow and restricted to a small number of institutions.^45,46^ If improvements in integration were to exist it would allow easier avenues for institutional monitoring of genetic testing. These data could point to areas of over-or under-testing that would help guide directions for potential improvements in utilization and efficacy.^47^ Successful direct EHR integration for genetic testing results or alternative approaches that extract information from PDFs of testing reports or that use large-language models to retrieve test results from unstructured clinical notes will be required to efficiently curate this information for clinical and research uses.

Our study has several key limitations. First, this work was conducted at a single academic medical center with a substantial clinical genetic footprint. Genetic testing trends and patterns may differ across institutions and clinical specialties, such that our results may not completely generalize to other locales. Second, we identified testing by indexing medical records for keywords, including gene names and genetic testing companies. This process may have missed genetic testing that was not recorded in the clinical notes and/or may have biased our dataset against negative tests. Third, our dataset is restricted to germline testing excluding testing for pharmacogenomics, fetal DNA, and tumor testing. We anticipate that these forms of testing could be incorporated using a slight modified approach.

In their current unstructured state, clinical genetic test results represent a vast and largely untapped resource. Structured test results can be used operationally to measure and monitor genetic test utilization and clinical utility. This dataset can be leveraged for research to improve understanding of how genetic diseases present in the EHR for improved detection and recognition. As clinical genetic testing becomes an ordinary part of health care, it has become necessary to capture testing data in a computable format to improve genetic testing and diagnostics in the future.

## Acknowledgements

This study was supported by a grant from the National Human Genome Research Institute (HG012657). Figure 1 was based on an example from the R Graph Gallery (https://r-graph-gallery.com) from Gilbert Fontana.

## Author contributions

LB conceived of the study and participated in all aspects of its execution

RJT Advised on the annotation of genetic disorders and panel types and assisted in the drafting and editing of the manuscript

BAS Assessed with the interpretation of results and assisted in the drafting and editing of the manuscript

JAP Advised on the content of the manuscript and provided comments.

WWS Advised on the content of the manuscript and provided comments. GH Advised on the content of the manuscript and provided comments.

JFP Assessed with the conception and implementation of the study and provided feedback on the manuscript. DMR participated in the design of the study and all aspects of assembling the manuscript.

## Data availability

We are unable to make the CGdb a publicly available resource due to privacy concerns. However, all resources used to conduct the study, including the extraction of genetic data from the EHR and all analyses, are available in the supplementary tables and files of this manuscript. The phecodeX mapping is available on our public website, phewascatalog.org.

