## Supplementary Figures for "Expansion of clinical genetic testing since the completion of the human genome project"

**Figure S1. *Percentage of genetic testing in structured versus unstructured clinical notes*.** The teal bars indicate the percentage of genetic testing results stratified by EHR source. Dark teal bars indicate tests that were available in structured reports (i.e., pathology reports or templated text embedded in clinical notes). Light teal bars indicate testing results that were recorded in clinical notes as narrative text.

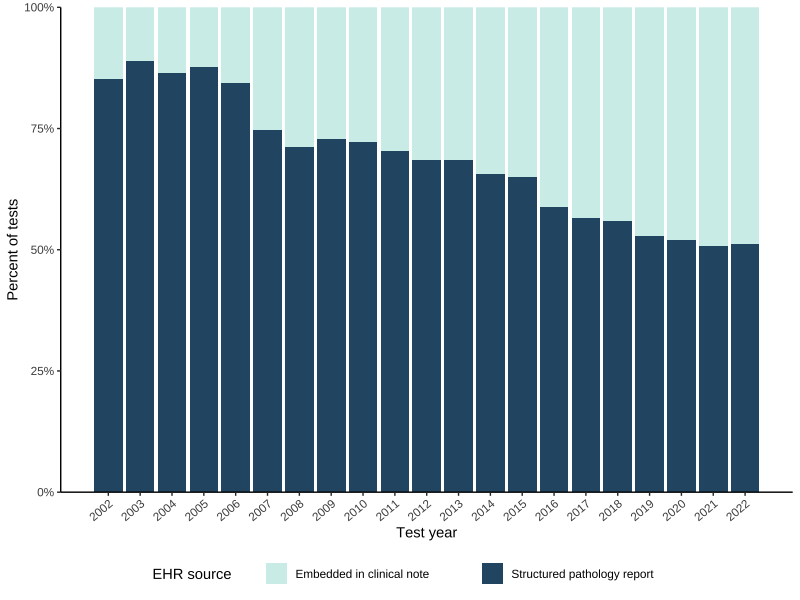

**Figure S2. *The fraction of patients seen each year with genetic testing and diagnoses*.** The solid lines show the percentage of patients receiving is indicated by the y-axis. The denominator of this percentage was defined as the number of patients seen at VUMC in the year indicated by the x-axis. The numerator was defined as the number of patients seen at VUMC that year who had genetic testing recorded in their EHR that year or any year previously. The dashed lines show the number of patients with a genetically confirmed diagnosis or risk factor in their EHR. The yellow line indicates all patients in the cohort. The purple and pink lines are stratified by adult (age >18 at time of visit) and pediatric (age<=18 at time of visit).

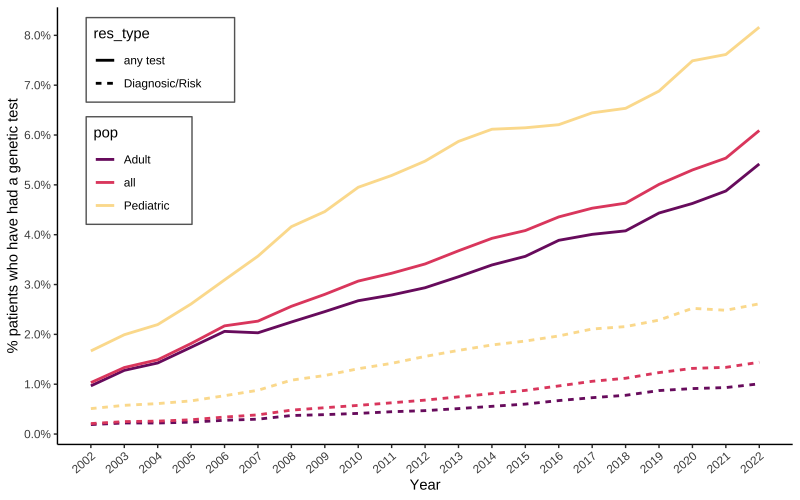

**Figure S3. Number of patients receiving tests each year, non-cumulative. (A)** The x-axis shows the number of new genetic tests introduced into the EHR per year, stratified by test type. **(B)** Same as plot A, except stratified by indication. In both plots, the y-axis indicates the number patients tested each year. Patients are counted only once per test type/indication and year.

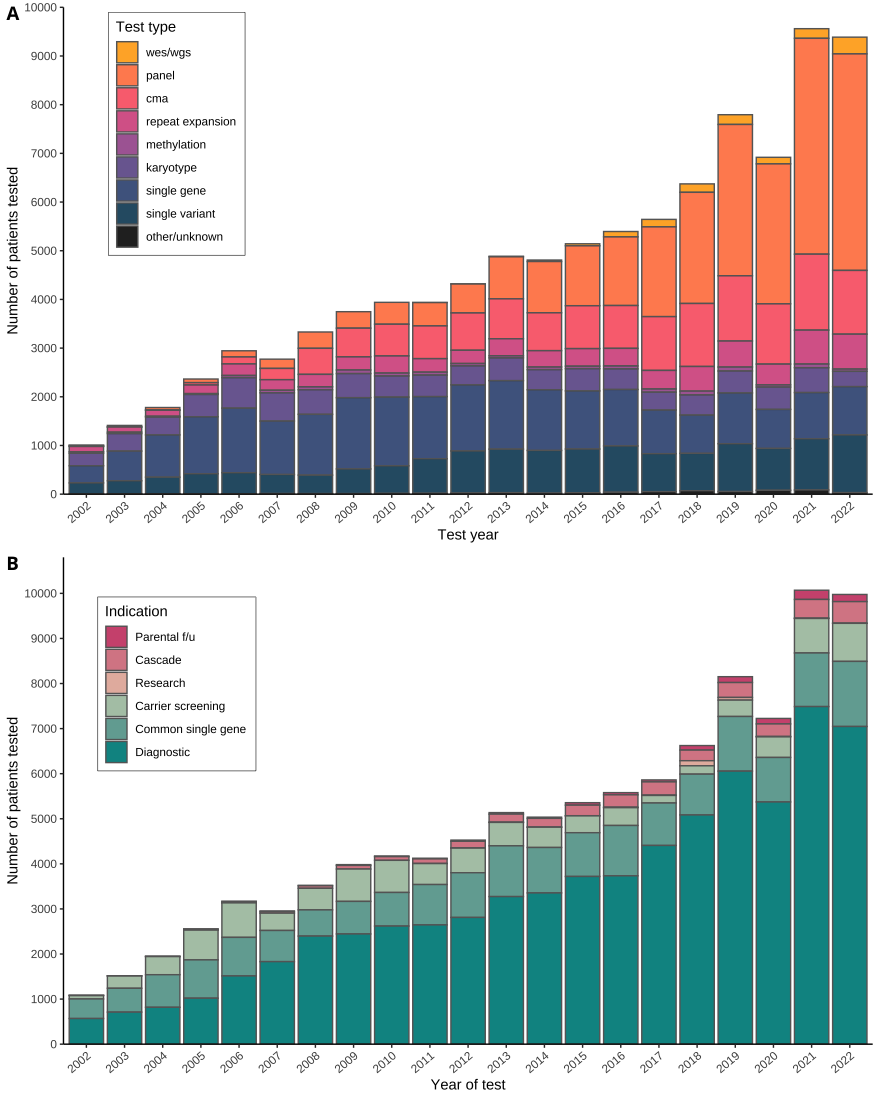

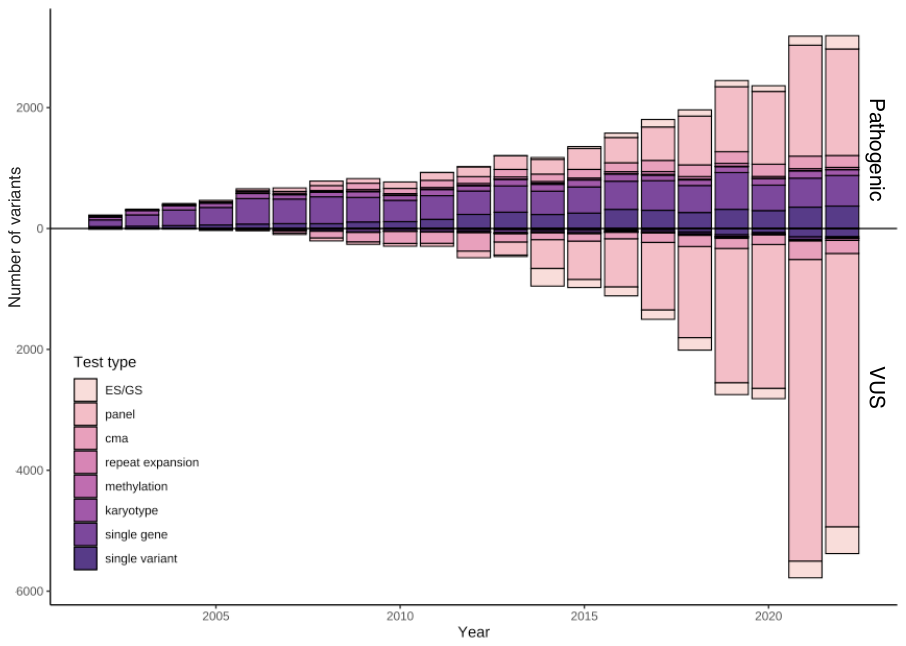

**Figure S4. The growth of VUS and differences in diagnostic vs. inconclusive tests across test types and disease categories**. The y-axis shows the number of variants recorded in the EHR per year. Counts for pathogenic variants are shown in vertical bars going upward and VUS counts are shown in bars going downward. Colors indicate tests type.

**Figure S5. Diagnostic vs. inconclusive tests across test type and panel disease categories**.

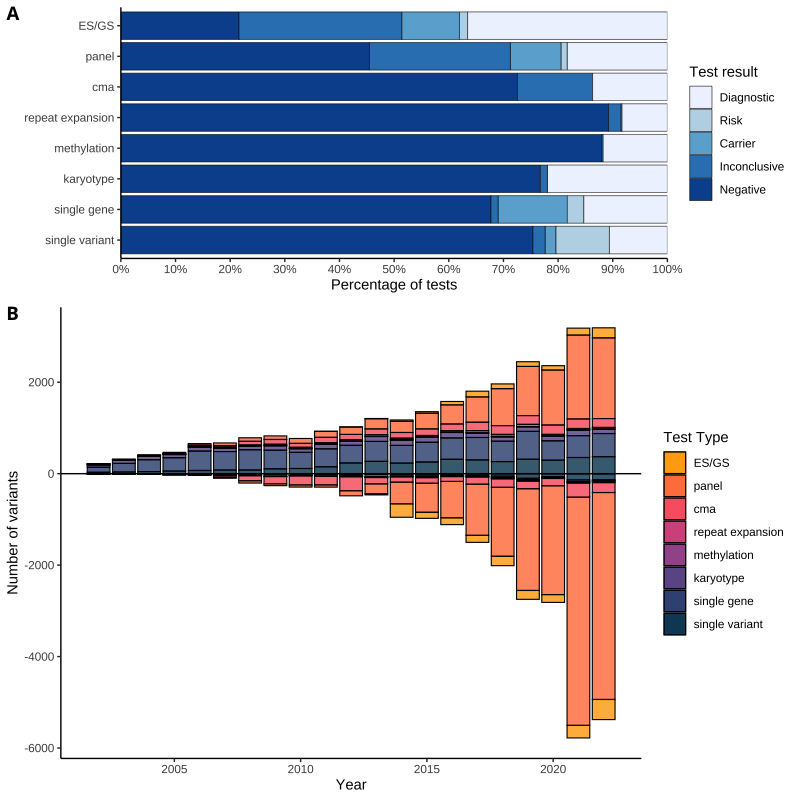

**Figure S6. Diagnostic vs. inconclusive tests across panel disease categories**. This plot shows test results for diagnostic panel testing for specific disease categories as well as ES/GS. All test disease categories with at least 100 instances and shown in the plot. The vertical dashed line shows the mean diagnostic yield of all panel and ES/GS tests.

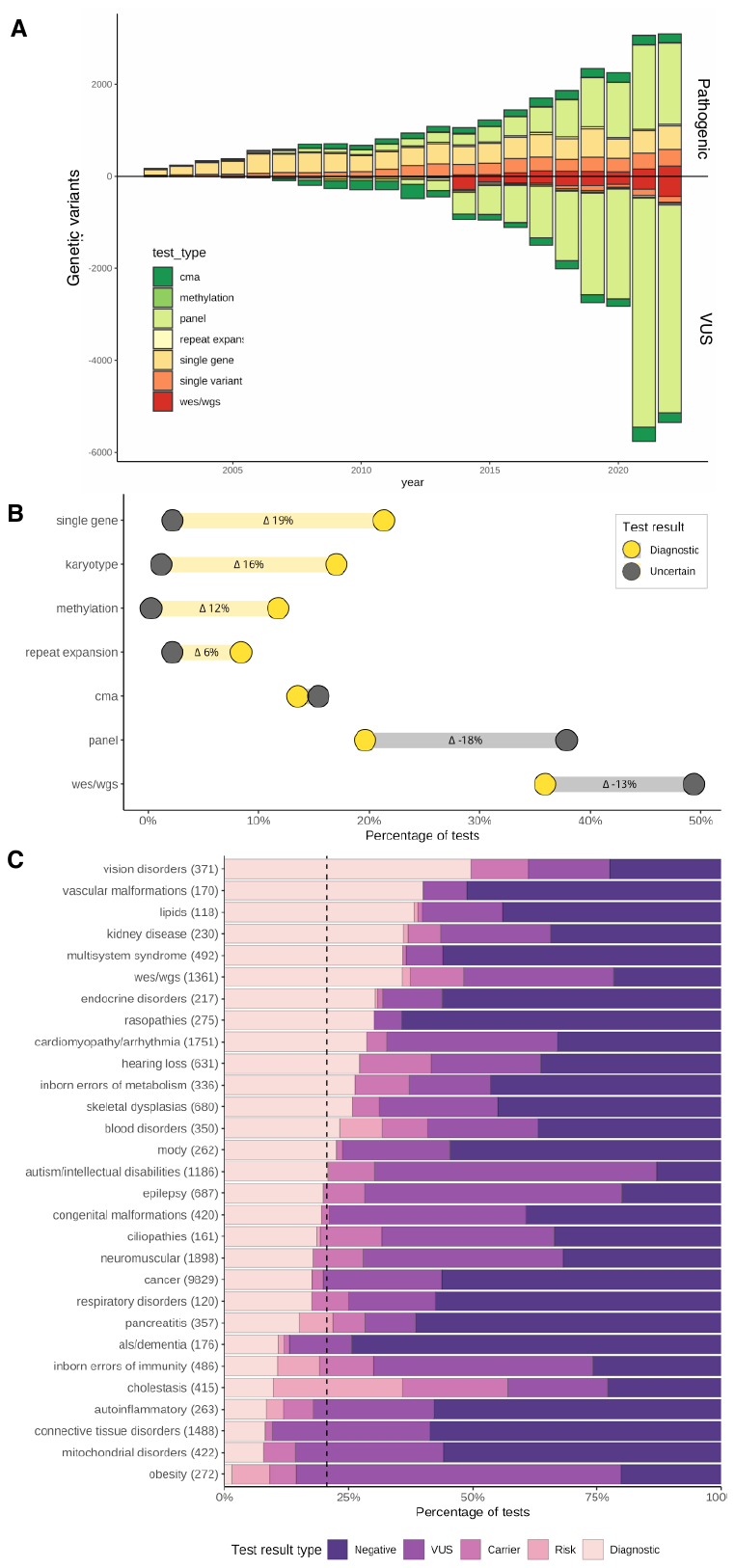

**Figure S7. Testing rate among patients visiting outpatient clinics in 2022**. The x-axis shows the fraction of patients with any genetic testing in their EHR, stratified by clinic type. The denominator of each bar includes individuals with an office and/or telemedicine visit to the specific clinic type.

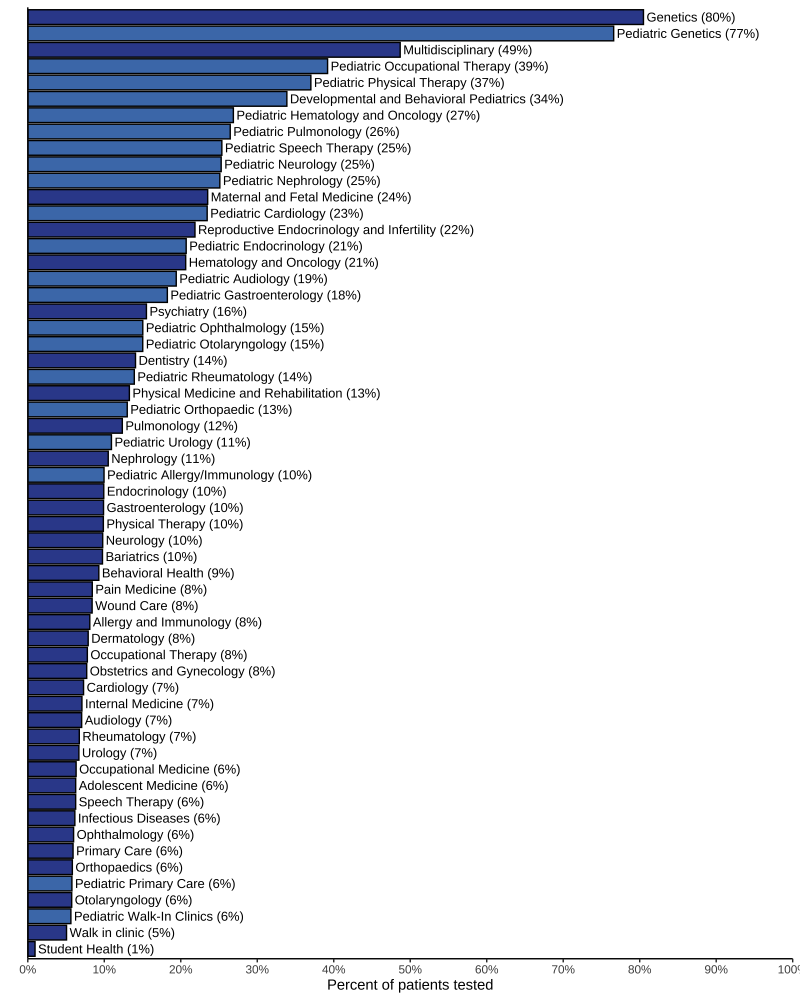

**Figure S9. Genetic attributable fraction (GAF).** The x-axis indicates the proportion of cases in the population of patients seen in 2022 that are attributable to a genetic diagnosis. Included in the figure are phenotypes with >4% GAF after pruning parent/child phenotypes. Phecode labels with a “*” indicate phenotypes only supported by ICD-10 codes.

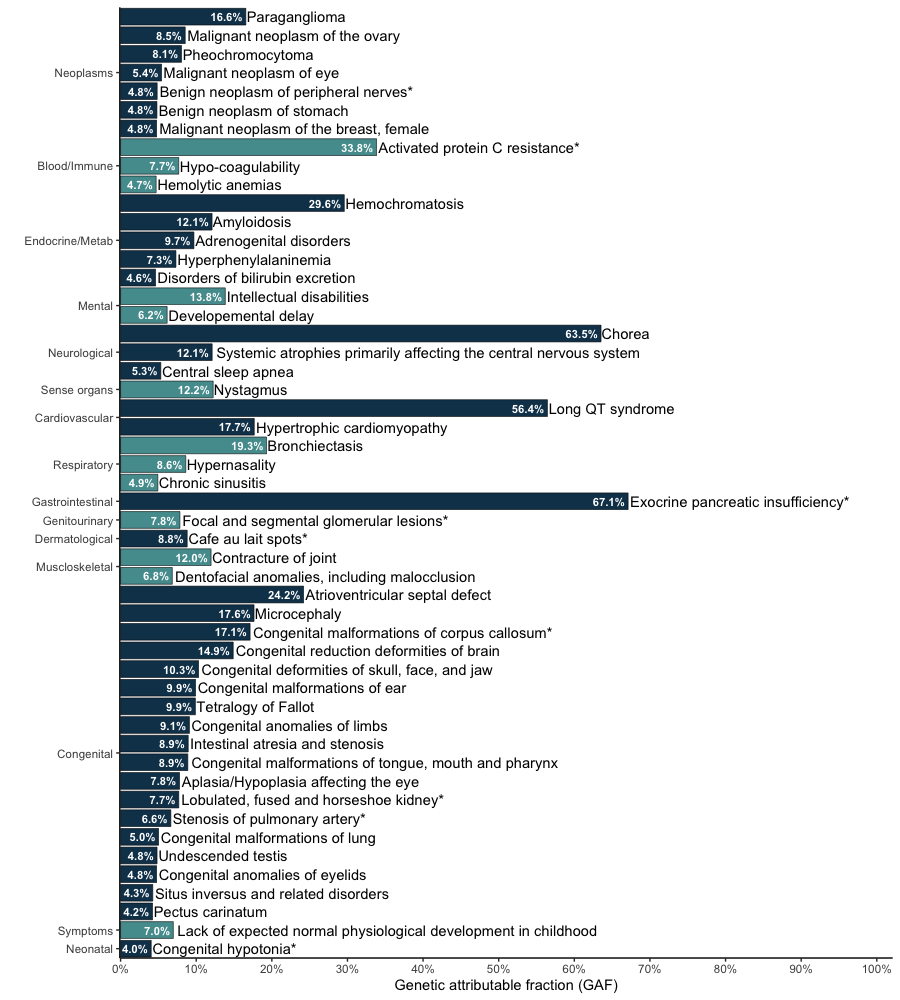
